# SARS-CoV-2 RNA and antibody detection in human milk from a prospective multicenter study in Spain

**DOI:** 10.1101/2021.05.06.21256766

**Authors:** Christine Bäuerl, Walter Randazzo, Gloria Sánchez, Marta Selma-Royo, Elia Garcia-Verdevio, Laura Martínez-Rodríguez, Anna Parra-Llorca, Carles Lerin, Victoria Fumadó, Francesca Crovetto, Fátima Crispi, Francisco J Pérez-Cano, Gerardo Rodríguez, Gema Ruíz-Redondo, Cristina Campoy, Cecilia Martínez-Costa, Maria Carmen Collado, on behalf of MilkCORONA study team

**Author notes:** Correspondence to: MCC, Institute of Agrochemistry and Food Technology, Spanish National Research Council (IATA-CSIC), Department of Biotechnology. Av. Agustin Escardino 7, 46980 Valencia, Spain. shared senior authorship (Cecilia Martínez-Costa, Maria Carmen Collado).

## Abstract

**Background:** During the COVID-19 pandemic in 2020, breastfeeding in women positive for SARS-CoV-2 was compromised due to contradictory data regarding potential viral transmission. However, growing evidence confirms the relevant role of breast milk in providing passive immunity by generating and transmitting specific antibodies against the virus. Thus, our study aimed to develop and validate a specific protocol to detect SARS-CoV-2 in breast milk matrix as well as to determine the impact of maternal SARS-CoV-2 infection on presence, concentration, and persistence of specific SARS-CoV-2 antibodies.

**Study design/Methods:** A prospective multicenter longitudinal study in Spain was carried out from April to December 2020. A total of 60 mothers with SARS-CoV-2 infection and/or recovered from COVID-19 were included (n=52 PCR-diagnosed and n=8 seropositive). Data from maternal-infant clinical records and symptomatology were collected. A specific protocol was validated to detect SARS-CoV-2 RNA in breast milk, targeting the N1 region of the nucleocapsid gene and the envelope (E) gene. Presence and levels of SARS-CoV-2 specific immunoglobulins (Igs) -IgA, IgG, and IgM-in breast milk samples from COVID-19 patients and from 13 women before the pandemic were also evaluated.

**Results:** All breast milk samples showed negative results for SARS-CoV-2 RNA presence. We observed high intra- and inter-individual variability in the antibody response to the receptor-binding domain (RBD) of the SARS-CoV-2 spike protein for each of the three isotypes IgA, IgM and IgG. Protease domain (MPro) antibodies were also detected in milk. In general, 82.9 % of the milk samples were positive for at least one of the three antibody isotypes, being 52.86 % of those positive for all three Igs. Positivity rate for IgA was relatively stable over time (65.2 – 87.5 %), whereas it raised continuously for IgG (47.8 % the first ten days to 87.5 % from day 41 up to day 206 post-PCR confirmation).

**Conclusions:** Considering the lack of evidence for SARS-CoV-2 transmission through breast milk, our study confirms the safety of breastfeeding practices and highlights the relevance of virus-specific SARS-CoV-2 antibody transfer, that would provide passive immunity to breastfed infants and protect them against COVID-19 disease. This study provides crucial data to support official breastfeeding recommendations based on scientific evidence.

## INTRODUCTION

Breastfeeding is considered the gold standard for infant feeding and is of crucial importance in influencing both infant growth and development, as well as in preventing future diseases during adulthood. Breast milk is a biological, complex, and living food that adapts to the specific nutritional and immunological requirements of the developing infant. Compared with formula feeding, exclusive breastfeeding has been consistently associated with lower morbidity and mortality during the first year of life, as well as with reduced incidence and severity of infections and immune-related diseases. Epidemiological studies have demonstrated that breastfeeding decreases risk of viral and bacterial infections, allergies, and obesity in infants [1–4]. Due to its beneficial effects, international organizations including the World Health Organization (WHO), the American Academy of Pediatrics (AAP), and European Society for Pediatric Gastroenterology Hepatology and Nutrition (ESPGHAN) Committee on Nutrition recommend exclusive breastfeeding for the first six months of life, and continuing breastfeeding while complementary foods are introduced until 2 years of age or beyond [5]. The global pandemic of coronavirus disease 2019 (COVID-19), caused by the severe acute respiratory syndrome coronavirus 2 (SARS-CoV-2), has increased the concerns on its mother-to-child transmission, including breastfeeding route. Currently, the scientific debate on whether breast milk could be a transmission vehicle for SARS-CoV-2 is still open and data are controversial. Some studies reported presence of the virus in breast milk [6,7], although its potential for infection is unclear [8]. Other studies found no presence of the virus in breast milk [9–11]. In general, these studies showed several limitations, with the most relevant being the lack of targeted and validated protocols for viral detection in milk matrix. Furthermore, a rapid and strong antibody response is induced after SARS-CoV-2 maternal infection, with the subsequent accumulation of substantial amounts of specific neutralizing secretory IgA (sIgA) in breast milk [12]. Other studies have also reported presence of specific antibodies in milk [7,13]. However, several questions still remain unanswered, including the extend of the response and persistence of maternal antibodies in milk, and their potential protective role for infants. Furthermore, exposure to other coronavirus might produce antibodies cross-reactive to SARS-CoV-2 [14]. Under this scenario, our main objectives are 1) to provide scientific-base evidence of the specific and reliable detection of SARS-CoV-2 in human milk; and 2) to determine levels of reactive IgA, IgG, and IgM antibodies against the receptor binding domain (RBD) of the spike protein and to the main protease (MPro) of SARS-CoV-2 in human milk samples collected during the COVID-19 pandemic.

## Material and Methods

### Study population

This is a prospective observational, longitudinal, and multicenter study in mother-infant pairs with confirmed SARS-CoV-2 infection (ClinicalTrials.gov Identifier: NCT04768244). Participants were recruited from 7 health centers from different provinces in Spain (Valencia, Barcelona, Granada, and Zaragoza). Recruitment period was from April to December 2020. Participants were pregnant with intended breastfeeding and nursing women with positive PCR for SARS-CoV-2 in nasopharynge or presence of SARS-CoV-2 antibodies in serum determined in hospitals. Women were excluded when COVID-19 symptomatology required specific treatment and/or hospitalization in intensive care units. All participants received oral and written information about the study and written consent was obtained. All protocols performed in the study were in accordance with the ethical standards approved by the Ethical Committee of the Hospital Clínico Universitario of Valencia (Ref. 2020/133) and by the rest of the Ethical and Research’s Committees.

A control group of women not exposed to SARS-CoV-2 and from prepandemic time was included. Those women were randomly selected from the MAMI birth cohort in Spain [15] (ClinicalTrials.gov Identifier: NCT03552939). This protocol was approved by the Ethical Committee of the Hospital Clínico Universitario of Valencia (Ref. 2015/0024) and by the local Ethical Committee of Atención Primaria-Generalitat Valenciana (CEIC-APCV).

### Human milk collection and processing

Breast milk collection was performed following a standardized protocol described elsewhere [16]. In brief, breast skin was cleaned with water and soap and the first drops were discarded. Then, milk was collected either by use of a sterile pump or manually extracted. Samples were collected in sterile bottles to normalize collection among participants. Morning collection was recommendable. Finally, breast milk samples were immediately stored at −201⍰°C in deep freezers and sent to the hospital to be stored at −80°C until further analysis.

Whole milk was used for SARS-CoV-2 RNA detection. Whey milk samples were used for antibodies determination and were prepared as follows: samples were thawed and centrifuged at 14,000 rpm at 4 °C for 10 min to remove fat and the resulting supernatant was transferred into new tubes. Centrifugation was repeated twice to ensure removal of all cells and fat. Skimmed acellular milk was then aliquoted and frozen at −80 °C until use. Prepandemic control milk samples were stored at −80°C before processing exactly as described for COVID-19 milk samples.

### Validation of SARS-CoV-2 RNA extraction, detection, and quantification in breast milk samples

A manual column-based commercial kit (referred as MN) and an automated assisted method based on magnetic beads (referred as Max) were adapted following previous recommendations [17] and compared to assess their sensitivity for detecting viral particles in breast milk samples. Main modifications of providers’ official protocols included: 150 μL of whole breast milk were treated with Plant RNA Isolation Aid (Ambion, USA) prior to extraction with MN (Nucleospin RNA virus Kit, Macherey-Nagel GmbH & Co., Germany), while 300 μL of whole breast milk were used for nucleic acid isolation with Max (Maxwell^®^ RSC Instrument coupled with Maxwell RSC Pure Food GMO and authentication kit, Promega, Spain). RNA was finally eluted in 100 μL nuclease-free water in both extraction protocols.

Initially, to characterize the viral recovery of both methods, porcine epidemic diarrhoea virus (PEDV) strain CV777 (an enveloped virus member of the *Coronaviridae* family and surrogate for SARS-CoV-2), and also, mengovirus (MgV) vMC0 (CECT 100000, non-enveloped member of the *Picornaviridae* designated in the ISO 15216-1:2017 standard method as process control) were spiked in phosphate buffer solution (PBS) and also, in breast milk. Detection ratios and limits of detection (LoD_95%_ and LoD_50%_) were also characterized for both extraction methods by spiking serial dilutions of PEDV.

Finally, serially diluted viral suspensions of gamma irradiated SARS-CoV-2 (Bei Resources; NR-52287), and human coronavirus (HCoV) 229E (ATCC-VR740) were used to further define the analytical performances of Max extraction. Detection limits were calculated using the PODLOD calculation program v.9 according to [18].

Viral RNA detection was performed by reverse transcription polymerase chain reactions (RT-qPCR) using One Step PrimeScript™ RT-PCR Kit (Perfect Real Time) (Takara Bio, USA), targeting the N1 region of the nucleocapsid gene for SARS-CoV-2 [19], the membrane gene for HCoV 229-E [20] and PEDV [21], and the region from 110 to 209 nucleotides for MgV [22]. The human RNase P gene [19] was used as quality control parameter for extraction. Reaction mixes, thermal cycling conditions, sequences for primers and probes, and standard quantification curves are detailed elsewhere [17,21]. Those for HCoV 229-E detection are included as supplementary material (Figure S1). Genome copies (gc) were calculated by using standard curves of 10-fold serial dilutions of SARS-CoV-2 genomic RNA (ATCC VR-1986D) or HCoV 229-E, PEDV and MgV suspensions in quintuplicates.

All RT-qPCR assays were performed in duplicate on a LightCycler 480 instrument (Roche Diagnostics, Germany). Positive (genomic RNA), negative (nuclease-free water), and inhibition (either 10-fold diluted RNA or RP gene) controls were included in each assay.

### Breast milk SARS-CoV-2-specific antibody detection

Levels of antibodies directed to structural proteins as RBD of the SARS-CoV-2 spike protein and to non-structural viral proteins as cysteine-like protease, also known as the main protease (Mpro) or 3CLpro, were analyzed.

RBD-specific antibodies were determined using a previously published and validated ELISA protocol for use in human plasma and serum samples [23,24], and was modified for its use in human milk samples. RBD protein was produced under HHSN272201400008C and obtained through BEI Resources, NIAID, NIH: Spike Glycoprotein RBD from SARS-CoV-2, Wuhan-Hu-1 with C-Terminal Histidine Tag, Recombinant from HEK293T Cells, NR-52946. Briefly, 96-well ELISA immunoplates (Costar) were coated with RBD protein at 2 µg/mL and incubated at 4 °C overnight. Coated plates were blocked in 3 % (w/v) milk powder in PBS containing 0.1 % Tween 20 (PBS-T) for 1 h. Then, 4-fold dilution of samples in 1 % (w/v) milk powder in PBS-T were added, incubated for 2 h at room temperature and washed with PBS-T before addition of horseradish peroxidase-conjugated secondary antibodies. For detection of the different antibody isotypes, anti-human IgA (α-chain-specific) HRP antibody (Thermo-Fisher Scientific; A18781; 1:6.000), anti-human IgM (μ-chain-specific) HRP antibody (Sigma-Aldrich; A0420; 1:4.000), and anti-human IgG (Fc specific) HRP antibody (Sigma-Aldrich; A0170; 1:4.000) were used and incubated for 1 h in 1 % (w/v) milk powder in PBS-T. Bound antigen-specific antibodies were detected with 100 μL 3,3⍰,5,5⍰-Tetramethylbenzidine (TMB) and reactions were stopped with 50 μL of 2M sulfuric acid. Absorbance at 450 nm was read in a ClarioStar (BMG Labtech) microplate reader using the path length correction mode. For detection of MPro-reactive antibodies, a commercial ELISA Kit (ImmunoStep, Salamanca, Spain) was used. Samples were incubated 1:4 diluted, and remaining steps of the protocol were performed according to manufacturer’s instructions. For ELISA studies, milk samples were considered positive when OD values from undiluted samples exceeded the positive cut-off values for each assay and isotype calculated from prepandemic control samples and defined as the mean +/-two standard deviations. Values from dilution curves were used for determining the area under the curve (AUC) to get a better quantitative impression between COVID-19 and control group. Endpoint titers were calculated from log-transformed titration curves using 4-parameter non-linear regression function in GraphPad Prism 8.0 and the positive cut-off values obtained from the prepandemic control group for each antigen and isotype.

### Breast milk Total IgA quantification

Total IgA, including secretory IgA (sIgA), was measured in whey milk using a sandwich ELISA quantitation kit from Bethyl Laboratories (Montgomery, TX) following the manufacturer’s instructions as previously detailed [25]. Briefly, an anti-human IgA antibody pre-adsorbed to the plate allowed to capture the IgA, which was later detected by the addition of a biotinylated detection antibody and streptavidin-conjugated horseradish peroxidase that catalyzed the colorimetric reaction with the chromogenic substrate TMB. All milk whey samples were analyzed at a 1:8,000 dilution. Data were expressed as mg/L of milk. Duplicate determinations were performed on each plate.

## Statistical analysis

Statistical analysis was performed in GraphPad Prism 8.0. After Shapiro-Wilk normality test, non-parametric t-test (Mann-Whitney) was used to detect significant differences between groups and Spearman correlation analysis to assess correlations between variables.

## Results

### Study population characteristics

A total of 60 mothers diagnosed with COVID-19 and 13 prepandemic were included in this study. The maternal demographic and clinical characteristics of the COVID pandemic period group (n=60) and the prepandemic group (n=13) are described in Table 1.

**Table 1:** Characteristics of the volunteers included in the study.

|  | COVID-19<br>(n=60) | Prepandemic Control<br>(n=13) | p-value |
| --- | --- | --- | --- |
| **Maternal characteristics:** |  |  |  |
| Age | 34.8 ± 4.6 <sup>1)</sup> | 33.8 ± 4.2 | 0.4827 <sup>a</sup> |
| Gestational age (weeks)* | 39.2 (38.1 – 40.6) <sup>2)</sup> | 39 (39.0 – 40.0) | 0.9631 <sup>a</sup> |
| Delivery mode, n (%) <sup>3)</sup> |  |  | 0.3062 <sup>c</sup> |
| Vaginal | 42 (76.4 %) | 8 (61.5 %) |  |
| Cesarean section | 13 (23.6 %) | 5 (38.5 %) |  |
| **Infant characteristics:** |  |  |  |
| Birth weight (g) | 3247 ± 519 <sup>4)</sup> | 3323 ± 475.7 | 0.6296 <sup>a</sup> |
| Birth length (cm) | 49.8 ± 2.4 <sup>5)</sup> | 50.5 ± 1.6 | 0.2958 <sup>a</sup> |
| Breastfeeding status <sup>6)</sup> |  |  | 0.7558 <sup>c</sup> |
| Exclusive | 35 (66.0 %) | 8 (61.5 %) |  |
| Mixed feeding | 18 (34.0 %) | 5 (38.5 %) |  |
| Gender <sup>7)</sup> |  |  | 0.5333 <sup>c</sup> |
| Male | 24 (44.4 %) | 4 (30.8 %) |  |
| Female | 30 (55.6 %) | 9 (69.2 %) |  |

\* Values are given as median and 25<sup>th</sup> and 75<sup>th</sup> percentile
<sup>a</sup> Unpaired t-test
<sup>b</sup> Mann-Whitney test
<sup>c</sup> Fisher's exact test (two-sided)
Missing data from <sup>1)</sup> 4 individuals; <sup>2)</sup> 10 individuals; <sup>3)</sup> 5 individuals; <sup>4)</sup> 8 individual; <sup>5)</sup> 14 individuals; <sup>6)</sup> 7 individuals;
<sup>7)</sup> 6 individuals

Among the 60 mothers, 52 were confirmed SARS-CoV-2 positive by PCR detection in nasopharyngeal swabs while the other 8 mothers were seropositive (IgG positive). Most PCR tests (38/52, 73·1%) were performed as part of routine surveillance before labor (Table 2). Three mothers had positive PCR-results postnatally (on days 9, 217, and 517 after delivery) and eight mothers had positive PCR results 72 h to 186 days prior to delivery. Out of the 8 seropositive mothers, 4 had a positive rapid antigen test within the month prior to delivery. Samples from two different time points during lactation were available for 12 of the women (<7 days and 15 days after delivery, approximately). Thus, a total of 72 breast milk samples from infected women and 13 from control women were included in the study.

**Table 2:** COVID-19 related data and sample characteristics.

| **COVID-19 diagnosis:** | **n=60** |
| --- | --- |
| Positive nasopharyngeal PCR confirmation | n=52 |
| confirmation 48 h before delivery | 38 |
| confirmation 48 h after delivery | 2 |
| confirmation > 48 h before delivery | 9 |
| confirmation > 48 h after delivery | 3 |
| Positive serology confirmation | n=8 |
| confirmation 48 h before/after delivery | 3 |
| confirmation > 48 h before delivery | 1 |
| confirmation > 48 h after delivery | 4 |
| **COVID-19 symptomatology** |  |
| Symptomatic prior or at enrollment | 22 |
| **Average time from COVID-19 diagnosis to breast milk sample collection** |  |
| Days post-PCR confirmation * | 15.0 (6.0 – 29.8) |
| Days post-serology confirmation * | 15.5 (2.0 – 40.5) |

\* Values are given as median and 25<sup>th</sup> and 75<sup>th</sup> percentile

Most of the women (n=38 (63·3%)) were asymptomatic and the rest reported only mild COVID-19 symptoms (pain, fatigue, or headache among others). No other effects or problems were reported. All neonates were negative for SARS-CoV-2 and were all in good health.

### Validation of SARS-CoV-2 RNA extraction and detection methods in breast milk

To optimize SARS-CoV-2 viral RNA detection in breast milk samples, an initial analytical comparison aimed to determine the recovery of PEDV and MgV from spiked prepandemic breast milk samples using manual (MN) and an automated (Max) extraction method. Compared to spiked PBS, PEDV and MgV were recovered at 30 % (27-33 %) and 132 % (94-188 %) when extracted from whole milk samples with MN. Better recoveries (1100 %) were observed for both viruses extracted by Max. No significant inhibitions due to the milk matrix were observed as depicted by cycle threshold (Ct) values of 10-fold diluted RNA or RP gene reactions.

By spiking PEDV serial dilutions in milk samples, we further defined the detection ratios and limits of detection (LoD_95%_ and LoD_50%_) (Table 3). Results demonstrated similar sensibility of both extraction methods being 10 and 13 PEDV gc/100μL the limit of detection with 95 % confidence for MN and Max extraction methods, respectively. These results suggested comparable analytical performances of both extraction methods for enveloped viruses, thus the latter was further characterized by using gamma inactivated SARS-CoV-2 and HCoV 229-E, along with PEDV and MgV, as the method intended to be used for screening breast milk samples from COVID-19. LoD_95%_ resulted in values as low as 36, 209, 13, and 7 gc/100μL, and LoD_50%_ in 8, 48, 3, 2 gc/100μL for SARS-CoV-2, HCoV 229-E, PEDV, and MgV, respectively (Figure 1). Based on these analytical results, the Max method was selected to screen the 72 breast milk samples for the presence of SARS-CoV-2 RNA. Targeting the N1 and E regions, all samples resulted negative for SARS-CoV-2 RNA presence. The RP gene used as quality control excluded false negative results (Cq= 27·98 ± 3·04). From 2 of the 72 samples, we used all the available volume for the PCR reaction and no remaining sample was available for following analyses. Thus, 70 milk samples from 60 women were available for further research.

**Table 3.** Concentrations, detection ratios and limits of detection (LoD_95%_ and LoD_50%_) characterizing the analytical performances of a manual commercial kit (MN) and an automated assisted method (Max) to extract porcine epidemic diarrhoea virus (PEDV) spiked in breast milk.

| Levels of inoculated PEDV (gc/100uL) | MN |  |  |  | Max |  |  |  |
| --- | --- | --- | --- | --- | --- | --- | --- | --- |
|  | Back calculated concentration (gc/100uL ± SD) | Detection ratio (+/total) | LoD<sub>95</sub> (gc/100 uL) | LoD<sub>50</sub> (gc/100 uL) | Back calculated concentration (gc/100uL ± SD) | Detection ratio (+/total) | LoD<sub>95</sub> (gc/100 uL) | LoD<sub>50</sub> (gc/100 uL) |
| 10e3 | 1348.84 ± 152.84 | 6/6 |  |  | 3137.92 ± 756.22 | 6/6 |  |  |
| 10e2 | 196.52 ± 10.10 | 6/6 |  |  | 575.86 ± 346.03 | 6/6 |  |  |
| 10e1 | 25.82 ± 5.03 | 6/6 | 10.0 | 2.96 | 92.94 ± 44.48 | 6/6 | 13.38 | 3.10 |
| 10e0 | 2.05 ± 0.25 | 2/6 |  |  | 16.39 ± 4.45 | 3/6 |  |  |
| 10e-1 | - | 0/6 |  |  | - | 0/6 |  |  |

**Figure 1.**
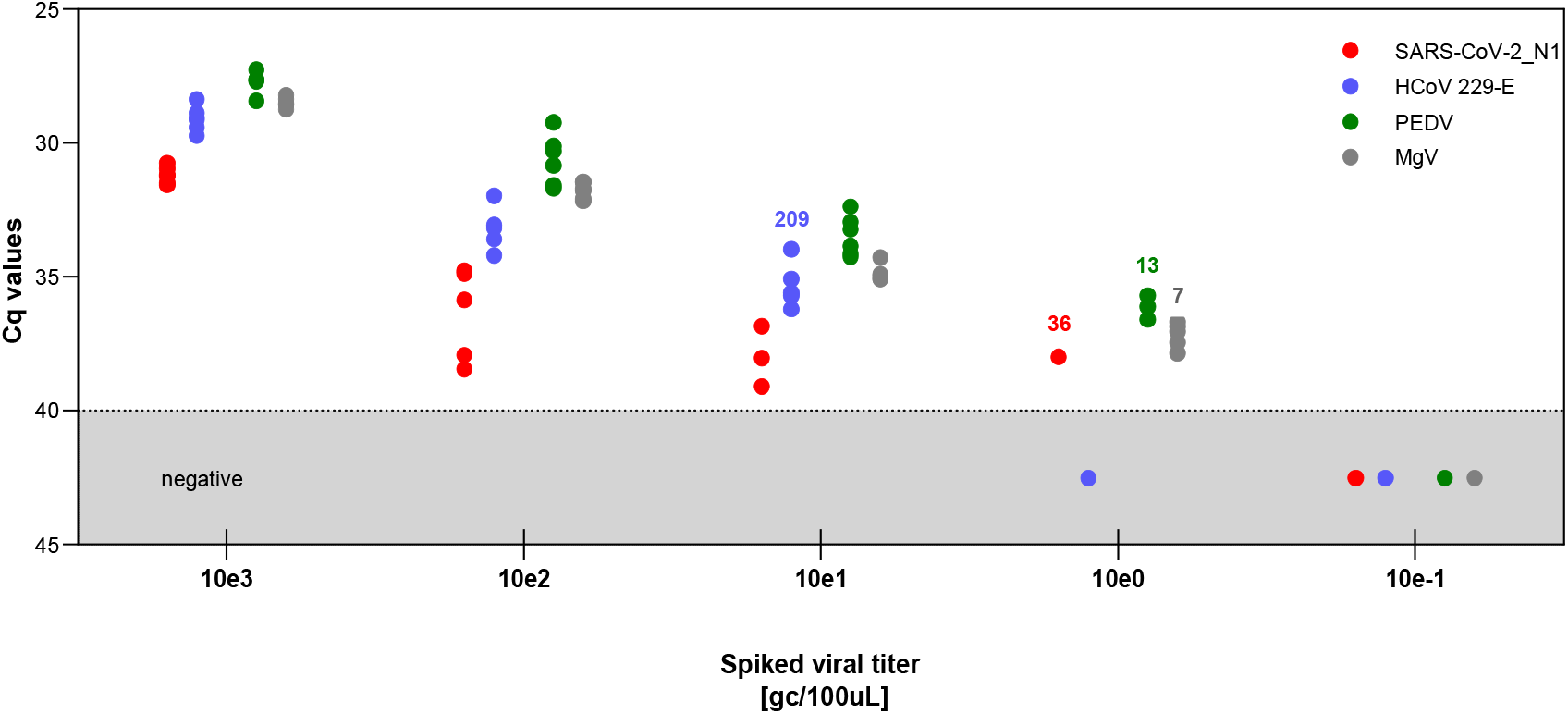
Distribution of cycle threshold values (Cq) characterizing the limits of detection of the automated assisted method (Max) to extract severe acute respiratory syndrome coronavirus 2 (SARS-CoV-2), human coronavirus 229E (HCoV 229-E), porcine epidemic diarrhea virus (PEDV), and mengovirus (MgV) spiked in breast milk. The numbers above viral series denotes the LoD_95%_ (gc/100uL) for each virus.

### SARS-CoV-2 reactive antibodies in breast milk

To characterize the antibody response against SARS-CoV-2 in breast milk, we first tested reactivity of IgA, IgM, and IgG to the RBD of the spike glycoprotein. Serial sample dilutions were performed and both AUC and endpoint titers were calculated. Prepandemic milk samples (n=13) served as controls and to determine positive cut-off values (Figure S2). Strong and significant reactivity was found for IgA, IgM, and IgG from COVID-19 infected/recovered breast milk and low levels of non-specific binding was observed for the prepandemic samples (Figure 2 a-c). When applying positive cut-off levels, 84·48 % (49/58) of the participating mothers provided milk samples that tested positive for the RBD antigen at least in one of the three antibody classes (Figure 2d) and in one of the repeated samples. When analyzing the 70 collected samples, 58 (82·9 %) were positive at least for one of the three antibody classes (IgA, IgM or IgG). Thirty-seven milk samples (52·9 %) were positive for all three Igs, whereas 12 samples (17·1 %) did not show reactivity against RBD for any of the three antibody classes (Figure 2e). In total, 51 samples were positive for IgA (72·9 %), 51 samples for IgM (72·9 %) and 45 samples for IgG (64·3 %). Among the samples that were positive for only one isotype, IgM reactive RBD antibodies exceeded cut-off levels in 4 samples, whereas for IgG and IgA only one sample showed reactivity against RBD for a unique isotype.

**Figure 2:**
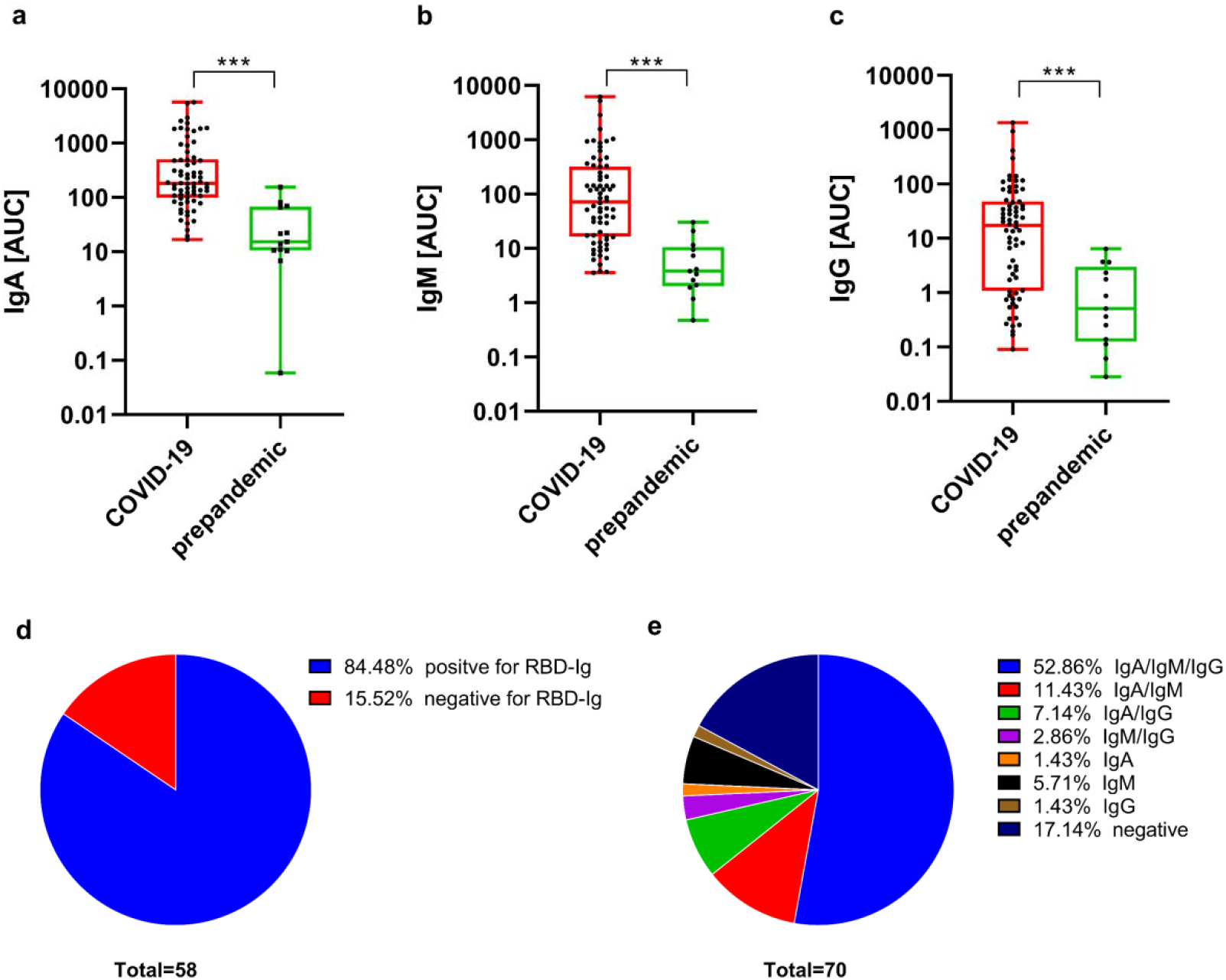
Reactivity of human milk samples from COVID-19 infected and/or recovered mothers and prepandemic controls to RBD antigen. AUC were calculated from titration curves for RBD-reactive (a) IgA, (b) IgM, and (c) IgG in order to get a better graphical impression. Asterisks show statistically significant differences between groups (^***^p < 0·0001) using the Mann–Whitney test (unpaired nonparametric test). (d) Proportion of human milk donors who had positive RBD-reactive Igs in at least one milk recollection point, and negative samples (e) Proportion of RBD-reactive positive and negative human milk samples subdivided according to different isotypes.

We corroborated our results using the MPro antigen, the main viral protease also known as 3CLPro as described elsewhere [26]. OD values of 1:4 diluted samples were grouped and COVID-19 infected and recovered donors still showed significantly higher reactivity to the MPro antigen (Figure 3). Noteworthy, positivity rate using this antigen decreased at this dilution rate from 67·6 % to 42·3% for IgA and from 64·2 % to 31·3 % for IgG.

**Figure 3:**
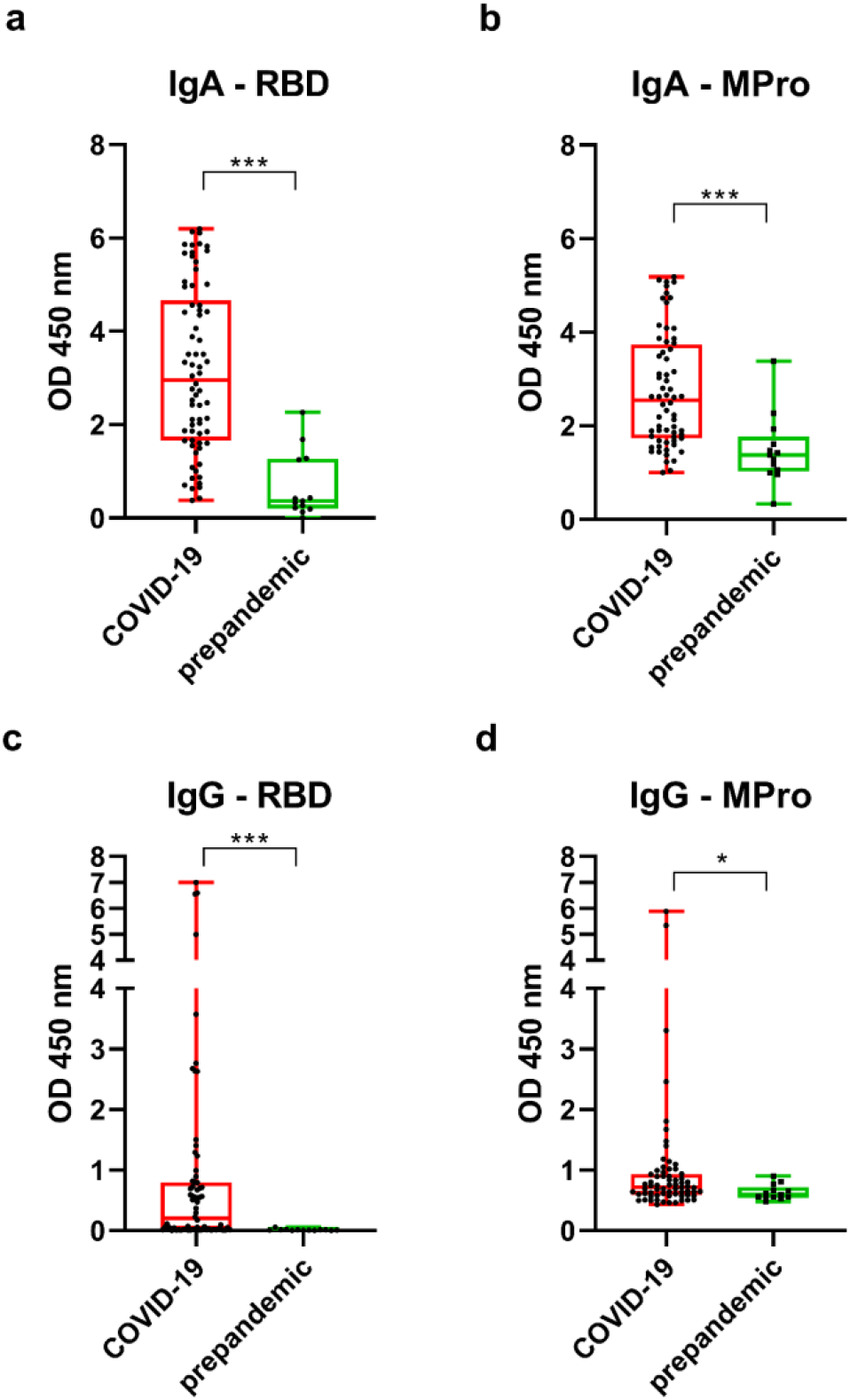
Samples from infected milk donors show significantly higher binding to SARS-Cov-2 antigens compared to prepandemic control. Grouped OD values of 1:4 diluted samples of RBD-(a) and MPro-(b) reactive IgA, and RBD-(c) and MPro-(d) reactive IgG, respectively. Asterisks show statistically significant differences between groups (^***^p<0·0001, ^*^p<0·05) using the Mann–Whitney test (unpaired nonparametric test).

Since most of the mothers in our study were confirmed positive for SARS-CoV-2 as part of routine surveillance prior to labor and therefore were asymptomatic, antibody response was analyzed as a function of time since PCR-confirmation (Table 4). Positivity rate for IgA was relatively stable over time (65·2–87·5 %). Most positive samples for IgM were detected when collected at 11-20 days after PCR confirmation (83·3 %), and then levels consistently declined to 62·5 %. IgG positivity rate continuously raised from 47·8 % to 87·5 % in samples collected from day 41 up to day 206 post-PCR confirmation. In symptomatic COVID-19 cases, RBD-specific IgA response was higher than in the asymptomatic group, although differences were not significant, and no changes were detected in virus-specific IgM and IgG (Figure S3).

**Table 4:** Positive samples and rates of virus-RBD specific antibodies versus days of positive PCR confirmation.

| **\* Days post-PCR** | **0-10d**<br>**(n=23)** | **11-20d**<br>**(n=18)** | **21-40d**<br>**(n=13)** | **41-206d**<br>**(n=8)** |
| --- | --- | --- | --- | --- |
| **IgA** | 15 (65.2 %) | 13 (72.2 %) | 9 (69.2 %) | 7 (87.5 %) |
| **IgM** | 18 (78.3 %) | 15 (83.3 %) | 7 (53.9 %) | 5 (62.5 %) |
| **IgG** | 11 (47.8 %) | 10 (55.6 %) | 10 (76.9 %) | 7 (87.5 %) |

\*Number of breast milk samples with positive results and percentage.

We compared endpoint titers of positive samples between the different antibody isotypes and observed that the magnitude of response was similar in all three Igs (Figure 4). Furthermore, all three Igs were significantly correlated to each other, particularly IgA and IgM (r= 0·7812, p<0·0001), but also IgA and IgG (r=0·6100, p<0·0001), and IgG and IgM (r=0·5708, p=0·0001).

**Figure 4:**
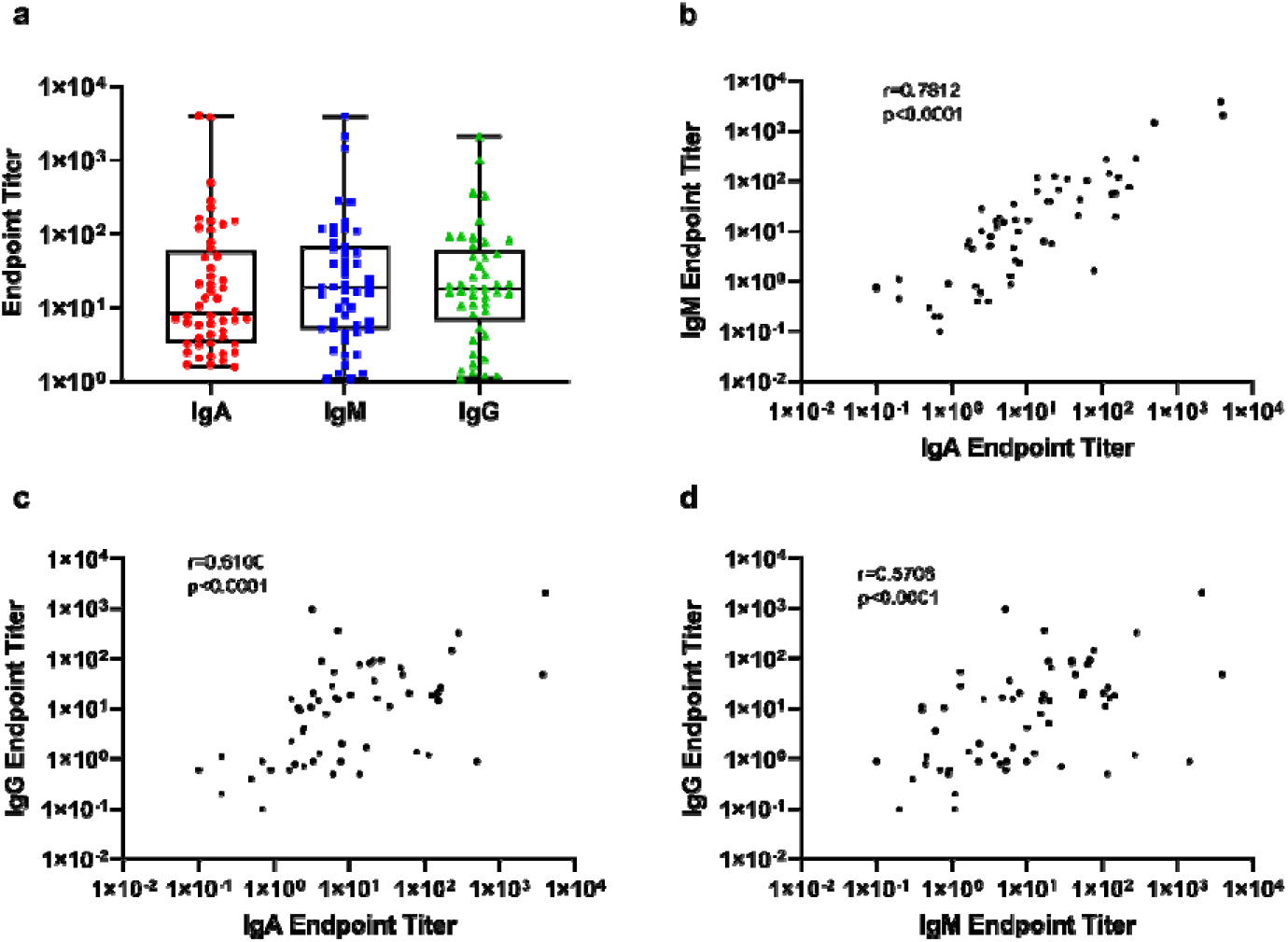
Endpoint titers and correlation analysis between RBD-specific binding of antibody subclasses. (a) Grouped endpoint titers of the three different isotypes. Spearmans’ correlation analysis of endpoint titers between (a) RBD-specific IgA and RBD-IgM, (b) RBD-IgA and RBD-IgG, and (c) RBD-IgM and RBD-IgG.

A positive correlation (r=0·5900, p=0·0001) was also observed between total IgA levels and SARS-CoV-2 specific antibody response (Figure 5a). In fact, total IgA levels were significantly higher in the COVID-19 group when compared to the prepandemic controls (Figure 5b) and could be part of the response to infection. In all the samples collected at different time points within the first 20 days after birth, we observed this generalized decrease of IgA and endpoint titers for RBD except in one mother, which exhibited low but raising antibody titers in breastmilk (Figure 6). In our samples, total IgA concentrations correlated negatively with lactational stage (r=-0·3357, p=0·0045), as well as with RBD-specific IgA (r=-0·3088, p=0·0093) and IgM (r=-0·4334, p=0·0002), while the RBD-specific IgG response was independent of the lactation stage. Furthermore, there was high inter- and intra-individual variability in the antibody response to the virus for each of the three isotypes (Figure 7). Since most of the donors were PCR-detected as part of birth COVID-19 protocol at hospitals, lactation stage and the days elapsed since PCR detection coincided in a narrow time fragment; in fact, for 40 of the positive tested samples in Figure 7a the difference was not more than 5 days between PCR-detection and birth. Nevertheless, some individuals showed a long-lasting antibody response and, in fact, two mothers which were positive PCR-tested 96 days (sample M39) and 186 days (sample M61) before birth showed virus-specific-antibody titers for all three Igs, having the highest levels for IgG. Regarding breastmilk samples from mothers confirmed with SARS-CoV-2 infection by serum serology (Figure 7b), seven out of the eight participants showed thoroughly positive antibody responses for all three antibody classes, except one sample that was tested negative for IgM. The sample that was negative for all three isotypes was from a mother with a COVID-19 infection confirmed by serological testing 226 days prior to sample collection for our study, with a 47-day old infant.

**Figure 5:**
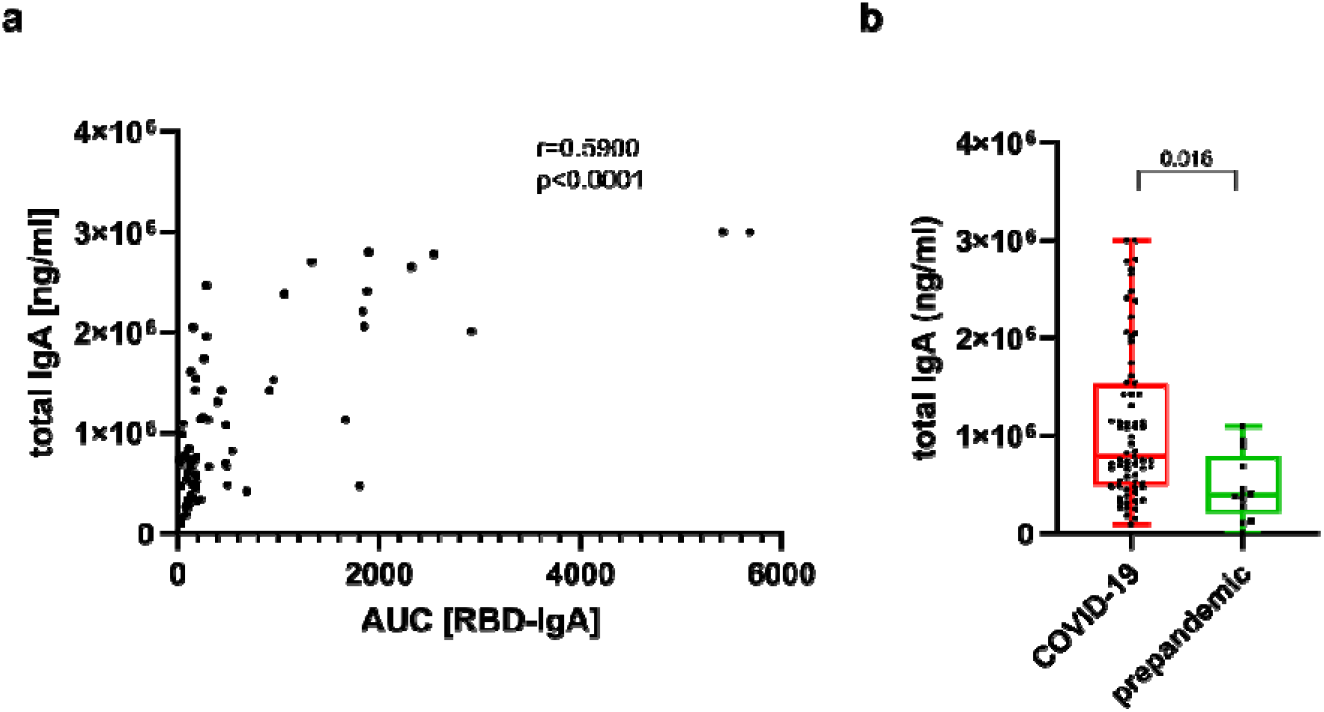
Total IgA concentration and virus specific antibody response. (a) Spearmans’ correlation analysis of total IgA concentration and virus specific IgA response expressed as AUC. (b) Total IgA in COVID-19 infected and recovered and prepandemic control samples.

**Figure 6:**
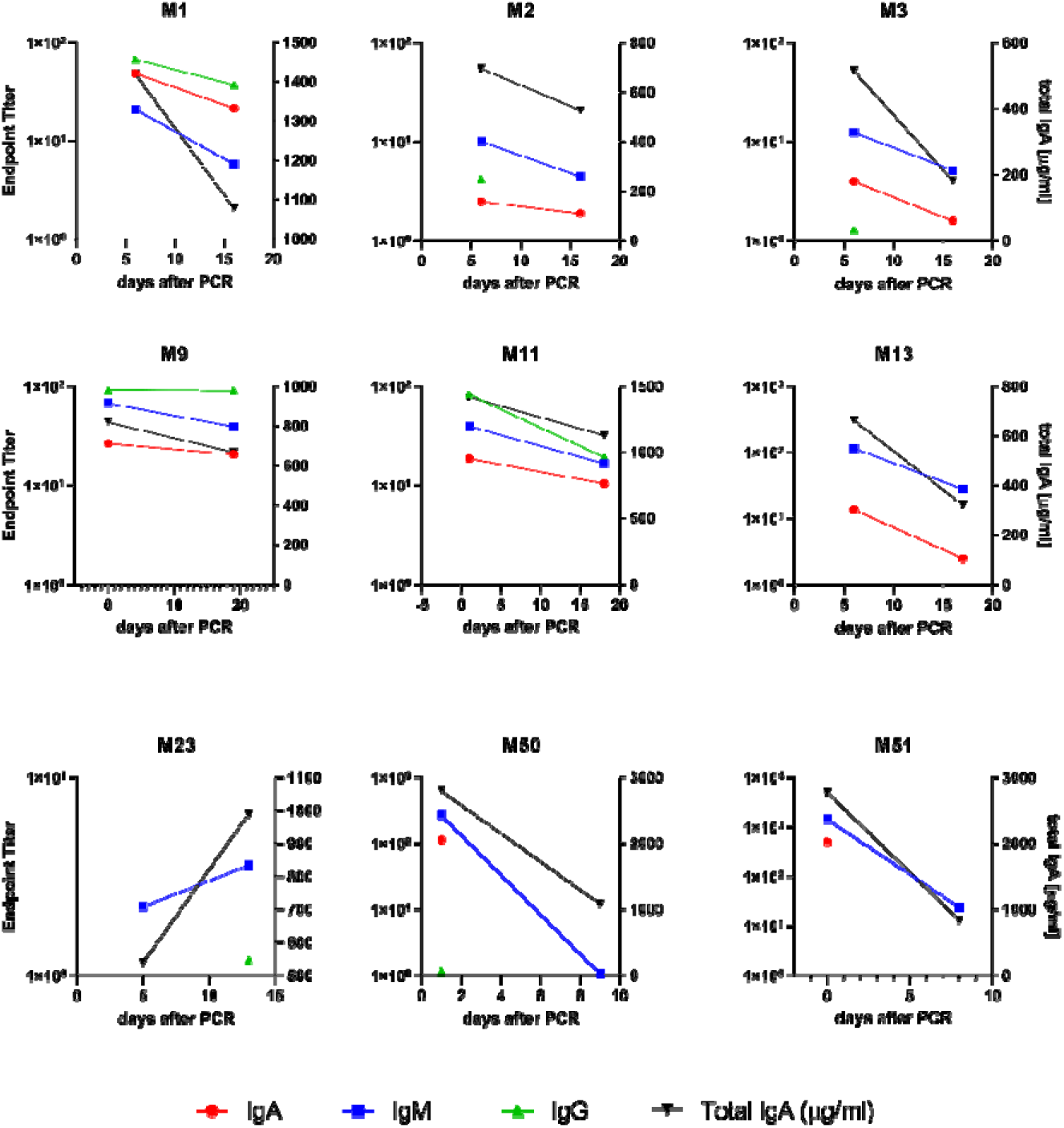
Temporal dynamic changes of endpoint titers for RBD-specific Igs. IgA (red circles), IgM (blue squares) and IgG (green triangules). All isotypes were tested for RBD binding in both time points, only positive endpoint titer are drawn in the graphs, the absence of data in a given time point indicates samples that were below the Cut-off values and considered negative.

**Figure 7:**
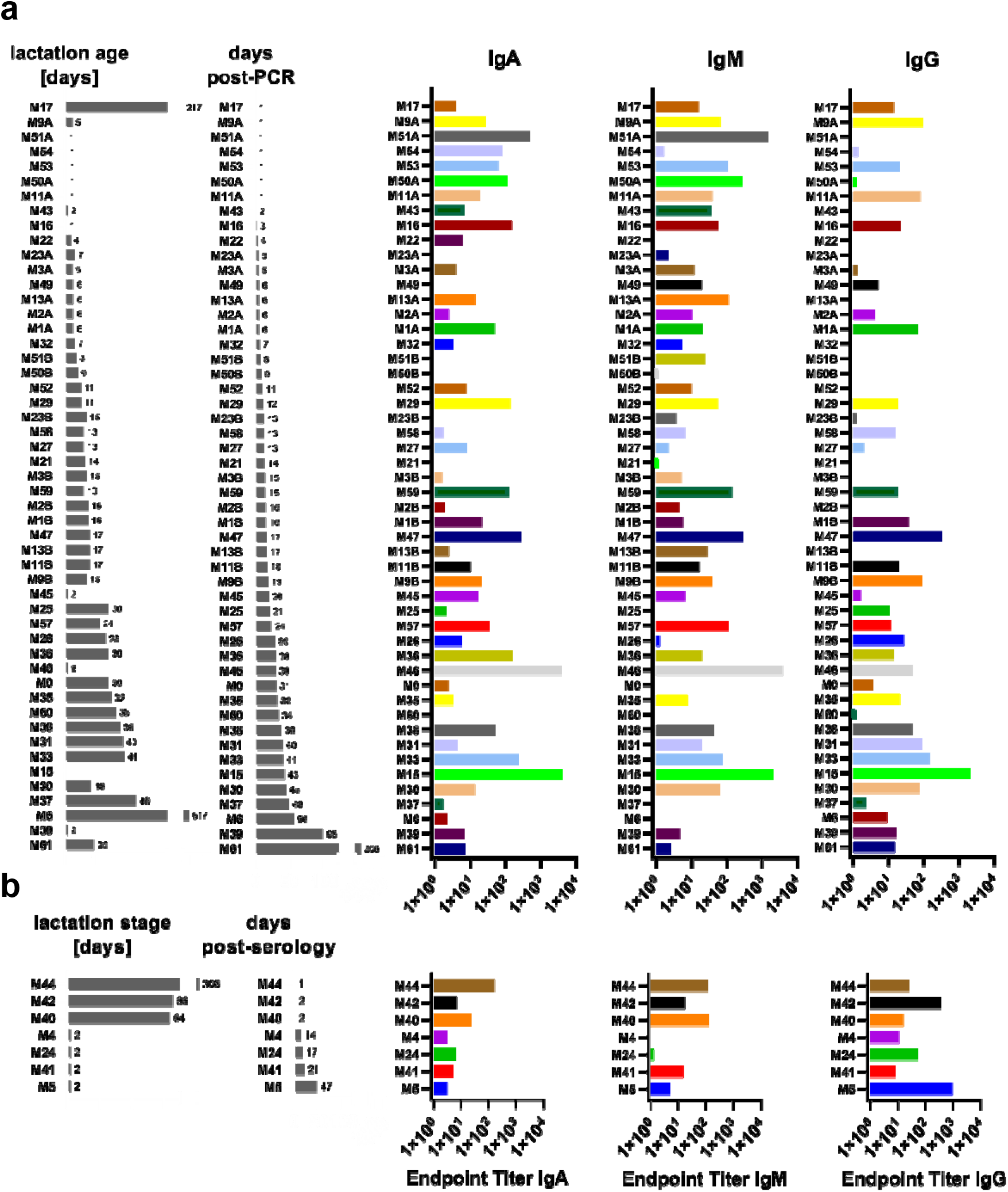
Individual endpoint titers of virus-specific antibodies in breast milk from SARS-Cov-2 infected and/or recovered mothers. (a) Endpoint titers of milk samples tested positive at least in one of the isotypes IgA, IgM and IgG are shown, ordered as days post-PCR. (B) Endpoint titers of milk samples tested positive at least in one of the isotypes IgA, IgM, and IgG are shown, ordered as days post-serology.

## Discussion

During the current COVID-19 pandemic, science has primarily focused on providing solutions and treatments against infection to reduce mortality. However, specific populations including pregnant and lactating mothers as well as infants have not been widely considered. Thus, there is a big gap in knowledge on maternal-infant health regarding COVID-19.

Breastfeeding is considered the most relevant postnatal link between mothers and infants, being the best source of nutrition with effects for infant health and development. However, the global COVID-19 pandemic and the lack of understanding of SARS-CoV-2 vertical transmission [27] have considerably reduced breastfeeding practice. Even infected mothers were recommended to temporarily separate from their infants [28].

In this prospective multicenter study, breast milk samples from women with COVID-19 were collected and tested for SARS-CoV-2 RNA presence by RT-PCRs validated for breast milk samples. It has been reported that DNA/RNA approaches could affect the detection of specific organisms in milk [29]; furthermore, milk matrix could affect the efficiency on nucleic acids recoveries [30,31] as we also identified. Thus, to facilitate the analysis, we chose whole milk to test for viral RNA presence. To provide alternative options for SARS-CoV-2 RNA detection, both a manual (MN) and an automated (Max) method were tested resulting in no significant differences between them. Moreover, viral RNA limit of detection in breast milk spiked with serial dilutions of a gamma inactivated SARS-CoV-2 stock were determined. The LoD95% resulted as low as 36 gc/100μL and the LoD50% 8 gc/100μL. These data are in line with the detection limit suggested by Chambers and colleagues where samples with >25 gc/100uL of SARS-CoV-2 RNA would be considered positive, [32], although higher limit of ca. 10^3-4^ gc/100uL was informed elsewhere [6]. Using Max extraction and SARS-CoV-2 detection by targeting the N1 and gene E regions, we demonstrated that SARS-CoV-2 RNA was not present in the breast milk samples tested.

Most of the bigger studies have been carried out in USA [7,8,32] and limited data are available for the European region. A study in an Italian cohort with 14 mother-infant pairs [33] showed that SARS-CoV-2 positive mothers do not transfer the virus during breastfeeding. Four neonates from this study were also positive, suggesting other potential infection routes. Another study in Germany with longitudinal samples from 2 women found some samples with positive signal for viral RNA [6]. A recent systematic review reported 37 articles analyzing breast milk samples on a total of 68 lactating mothers with COVID19 and SARS-CoV-2 RNA was detected in 9 of the samples (9/68, 13·2%) [34]. Available data in recent literature showed that around 2-6 % of milk samples would harbor viral RNA. A recent systematic review including 116 confirmed COVID-19 lactating women reported that the SARS-CoV-2 RNA detection in human milk was 2·16 % [35]. The biggest study up to now included study, with 110 women in USA (n=65 testing positive for SARS-CoV-2), showed that 6 % of the women had RNA presence of SARS-CoV-2 in milk, however no infectious viral particles were isolated by cell culture [8]. Despite this evidence, lactating women should follow the recommendations of wearing a mask, washing their hands during lactation, and avoiding droplets or aerosols that can infect the neonate. In our study, we have not detected SARS-CoV-2 RNA in any of the breast milk samples from participants in our study, contributing to the evidence that there is no vertical transmission during breastfeeding. Our study represents one of the biggest studies in Europe and the first in Spain. Currently, there is no evidence of SARS-CoV-2 transmission through breast milk. Regarding the potential limitations, we did not collect skin swabs to control for the potential presence of SARS-CoV-2 in the skin [7,33]. However, all milk samples as well as the infants were negative for SARS-CoV-2 presence. Thus, although there are still limited data, it is accepted that breastfeeding does not represent a vehicle for vertical transmission of SARS-CoV-2 [9]. There are still many open questions: when are SARS-CoV-2 antibodies produced after maternal infection, when can they be detected in breast milk, and how long do they persist. To cover some of these questions, we aimed to determine the presence of antibodies in breast milk samples from COVID-19 women and to compare these with milk samples collected prior to the pandemic as reference controls.

While different studies have reported presence of specific IgA antibodies against SARS-CoV-2 [7,12,13,36], limited information is available on IgG and IgM. Our results showed presence of anti-SARS-CoV-2 antibodies in milk, primarily IgA but also IgG and IgM targeting RBD. Furthermore, IgA- and IgG against non-structural MPro were analyzed for the first time in human milk samples. High intra- and inter-individual variability was observed in antibody presence and significant differences were observed for all three antibody classes when compared to prepandemic samples in agreement with previous data [26]. In our dataset we found that 82·9 % of the milk samples tested positive for the RBD antigen at least in one of the three antibody isotypes and 52·86 % were positive in all three Igs. Interestingly, 12 samples (17·14 %) did not show reactivity against the RBD region in any of the three antibody classes. Endpoint titers for all three antibody classes were in the same range and correlated significantly. We could not detect time-dependent quantitative differences in endpoint titers for the different antibody classes, due to high interindividual variability. However, we found a time-dependent increase from 47·8 % to 87·5 % in IgG-positive samples collected from day 41 up to day 206 post-PCR confirmation. These data are in line with findings in human serum samples from a Chinese cohort showing persistence of IgG up to 6 months in at least 90 % of confirmed cases [37]. To date, only a few studies have quantified virus-specific antibodies and compared them to prepandemic samples in order to establish cut-off values to discriminate between positive and negative samples [7,12,38], which allows the comparison of results between studies. We found similar proportion of positive samples for both IgA, from 72·9 % in our study to 80·0 % [7,12] and IgM, 72·9 % in our study compared to 55·0 % in the study reported by Peng and colleagues [38].

In this study, we also assessed presence of antibodies against other nonstructural viral proteins, specifically the viral cysteine-like protease, also known as 3CLPro or main viral protease (MPro), as described elsewhere [26]. Our data showed presence of anti-MPro IgA and IgG antibodies in milk samples from COVID-19 women compared to prepandemic samples, although the sensitivity was lower than when using the RBD antigen for detection of virus-specific antibodies. MPro is a viral antigen not exposed on the viral particle as this is the case of the spike protein, however, strong and similar reactivities were found for both MPro, RBD, and the nucleocapsid protein in serum and saliva samples [26]. Our study is the first using MPro for detection of SARS-Cov-2 antibodies in human milk, and the different reactivities to the different viral antigens, also in function of isotype, have been previously reported in human milk samples [7,33].

It is known that human milk contains a wide variety of Igs, with IgA representing the majority (80 to 90%). Secretory IgA (sIgA) is the predominant immunoglobulin class in human milk, representing about 90% of the total Igs present in human milk [39]. Our data showed positive associations between total IgA levels in milk and COVID-19 specific antibody responses. In fact, total IgA levels were significantly higher in the COVID-19 group when compared to the prepandemic controls, possibly due to the response to the infection. Total IgA concentrations in human milk change over time during the lactation period, with levels decreasing from colostrum to mature milk [40]. In this line, in samples collected at different time points within the first 20 days after birth, we observed this generalized decrease of IgA and endpoint titers for RBD, except in one mother, which noteworthy exhibited low but raising antibody titers in breast milk.

Our results are in agreement with previous data showing higher levels of antibodies against SARS-CoV-2 in milk compared to samples from women before the pandemic [36,41]. However, prepandemic samples showed some reactivity to SARS-CoV-2, which may be explained by cross-reaction with other seasonal coronavirus (HCoV) as previously reported [14]. It has been demonstrated that viral infections influence the antibody response in milk [42]. In addition, a recent study reported higher S1 + S2-reactive IgG in breast milk from women that had viral respiratory infection symptoms before pandemic than in milk from women without symptoms [36]. These data would partially explain the potential signal or cross-reactivity in breast milk samples before 2020.

Furthermore, SARS-CoV-2 IgA levels are higher in human breast milk than in maternal plasma while IgG and IgM levels are lower [43], suggesting potential active local production as reported for other virus including rotavirus [44]. We observed that women with COVID-19 symptoms showed slightly higher virus-specific IgA levels in milk compared to women with asymptomatic infection, although differences were not significant. Moreover, no differences in IgM or IgG levels were found, possibly due to minor COVID-19 symptoms (pain, headache, etc..) in this data set. Despite these observations, further analysis including a bigger sample size and different symptoms as well as severe COVID-19 infected donors are warranted. Regarding breastmilk samples from mothers confirmed COVID-19 by serum serology, only 1 out of 8 samples was negative for all three isotypes, possibly due do the time elapsed (226 days) between serological testing and sample collection.

In summary, our study demonstrates 1) absence of SARS-CoV-2 in breast milk from women with COVID-19; and 2) high inter- and intra-variability in SARS-CoV-2 antibody response. Women with a positive PCR or serology exhibited IgA, IgG, and IgM antibodies in breast milk not only against structural proteins like RBD but also against non-structural proteins like MPro. Presence of IgM in some samples suggests that breast milk might have a protective effect on newborns. Interestingly, positive associations between total IgA and specific antibodies (IgA, IgG and IGM) were observed, although their persistence and stability differed between mothers and antibody type. Our study endorses the safety of breastfeeding practices and highlights the potential relevance of virus-specific SARS-CoV-2 antibodies providing passive immunity to breastfeeding infants protecting them against COVID-19.

This study provides scientific evidence to support official recommendations stating breastfeeding safety during the COVID-19 pandemic, indicating that breastfeeding should be a priority with potential benefit for both mothers and neonates.

## Data Availability

data are available upon request

## Acknowledgements

We thank all the families who were involved in the study during this difficult time and in the middle of the COVID19 pandemic as well as the collaborators of the MilkCORONA (detailed below), which includes neonatologists, pediatricians, midwifes, nurses, research scientists, and computer/laboratory technicians. This study represents a cooperative effort between different groups in Spain.

## MilkCORONA Collaborators

i. Elena Crehuá-Gaudiza (Pediatrician, MD, PhD), Javier Estañ-Capell (Pediatrician, MD, PhD) from Pediatric Nutrition Research Group of INCLIVA Biomedical Research Institute of Valencia and Department of Pediatrics, Hospital Clínico Universitario, University of Valencia, Spain.
ii. Asuncion Obiol (midwife, PhD), Reyes Balanza (MD, PhD), Department of Gynecology and Obstetrics, Hospital Universitario Dr. Peset, Valencia, Spain
iii. Álvaro Solaz-García (RN), Inmaculada Lara-Cantón (MD), Health Research Institute La Fe, Neonatal Research Group, Spain and University and Polytechnic Hospital La Fe, Division of Neonatology, Valencia, Spain.
iv. Cristina Garcia (MsC), Institut de Recerca Sant Joan de Déu, Hospital Sant Joan de Déu, Barcelona, Spain.
v. María Ríos Barnés (pediatrician, MD) Infectious Diseases Unit. Pediatrics Department, Hospital Sant Joan de Déu, Barcelona, Spain.
vi. Sara Ruiz (MD, PhD), Marta Fabre (MD, PhD), Hospital Clínico Universitario Lozano Blesa, Zaragoza. Instituto de Investigación Sanitaria Aragón (IIS Aragón),
vii. Federico García-García (MD, PhD) Department of Microbiology. University Clinical Hospital San Cecilio, Granada and BioHealth Research Institute (Ibs), Granada, Health Sciences Technological Park, Granada, Spain.
viii. Maria José Rodríguez-Lagunas (PhD), Karla Río-Aigé (MsC), Physiology Section, Department of Biochemistry and Physiology, Faculty of Pharmacy and Food Science, University of Barcelona (UB), Barcelona, Spain and Nutrition & Food Safety Research Institute (INSA-UB), 08921 Santa Coloma de Gramenet, Spain.

## Contributions

Study concept and design: CMC and MCC. Drafting of the manuscript: CB, WR, CMC and MCC. Clinical aspects including recruitment of the participants, collection of samples and medical records: CMC, LMR, APLl, EGV, CL, VF, FC, FC, GR, CC, GRR. Sample management, processing, and analysis: CB, WR, MSR, GS, FJPC, MCC. All authors read and approved the final manuscript.

