## Supplementary material for "SARS-CoV-2 RNA and antibody detection in human milk from a prospective multicenter study in Spain"

### Slide 1
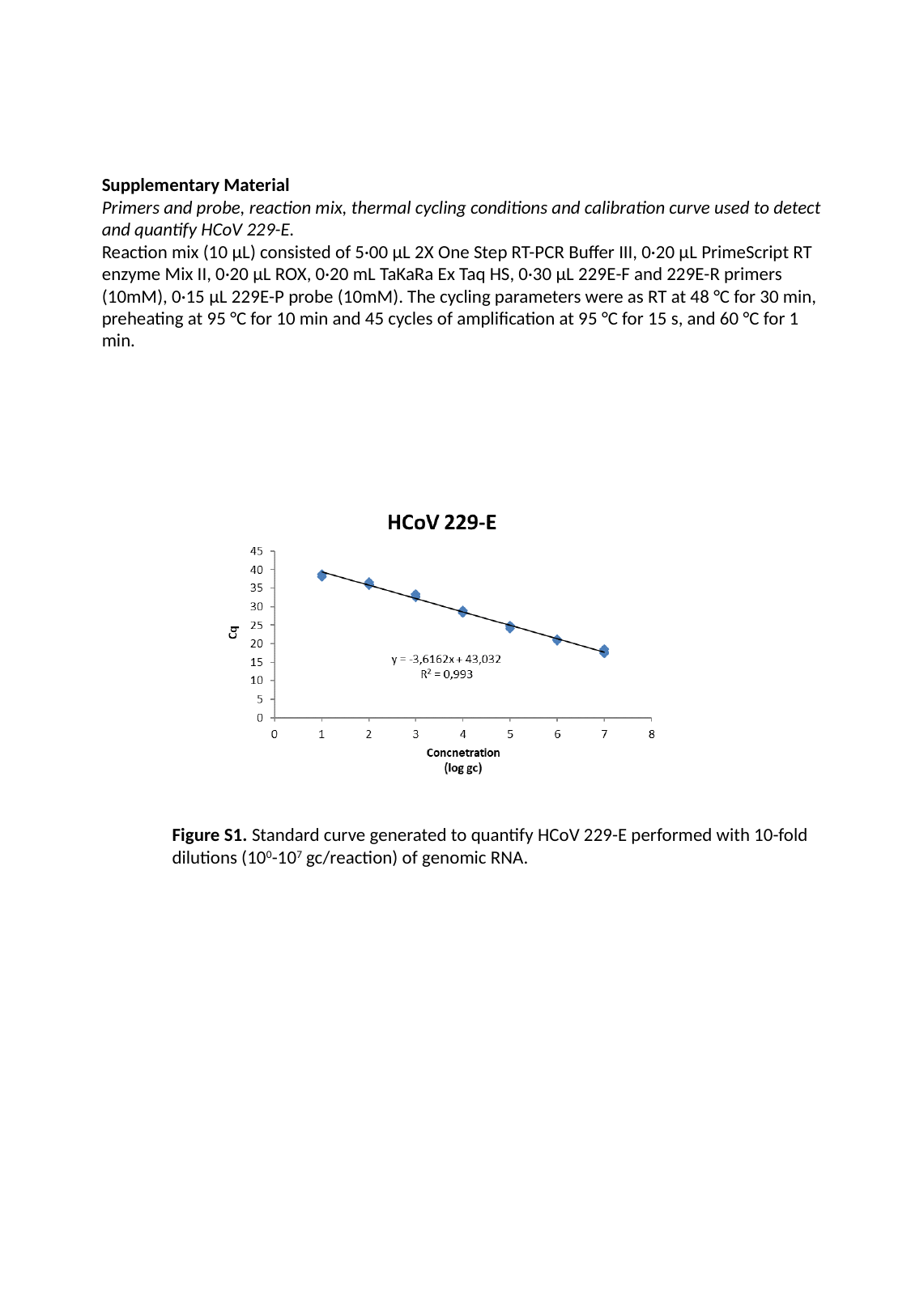

Supplementary Material
Primers and probe, reaction mix, thermal cycling conditions and calibration curve used to detect and quantify HCoV 229-E.
Reaction mix (10 µL) consisted of 5·00 µL 2X One Step RT-PCR Buffer III, 0·20 µL PrimeScript RT enzyme Mix II, 0·20 µL ROX, 0·20 mL TaKaRa Ex Taq HS, 0·30 µL 229E-F and 229E-R primers (10mM), 0·15 µL 229E-P probe (10mM). The cycling parameters were as RT at 48 °C for 30 min, preheating at 95 °C for 10 min and 45 cycles of amplification at 95 °C for 15 s, and 60 °C for 1 min.
Figure S1. Standard curve generated to quantify HCoV 229-E performed with 10-fold dilutions (10^0^-10^7^ gc/reaction) of genomic RNA.

### Slide 2
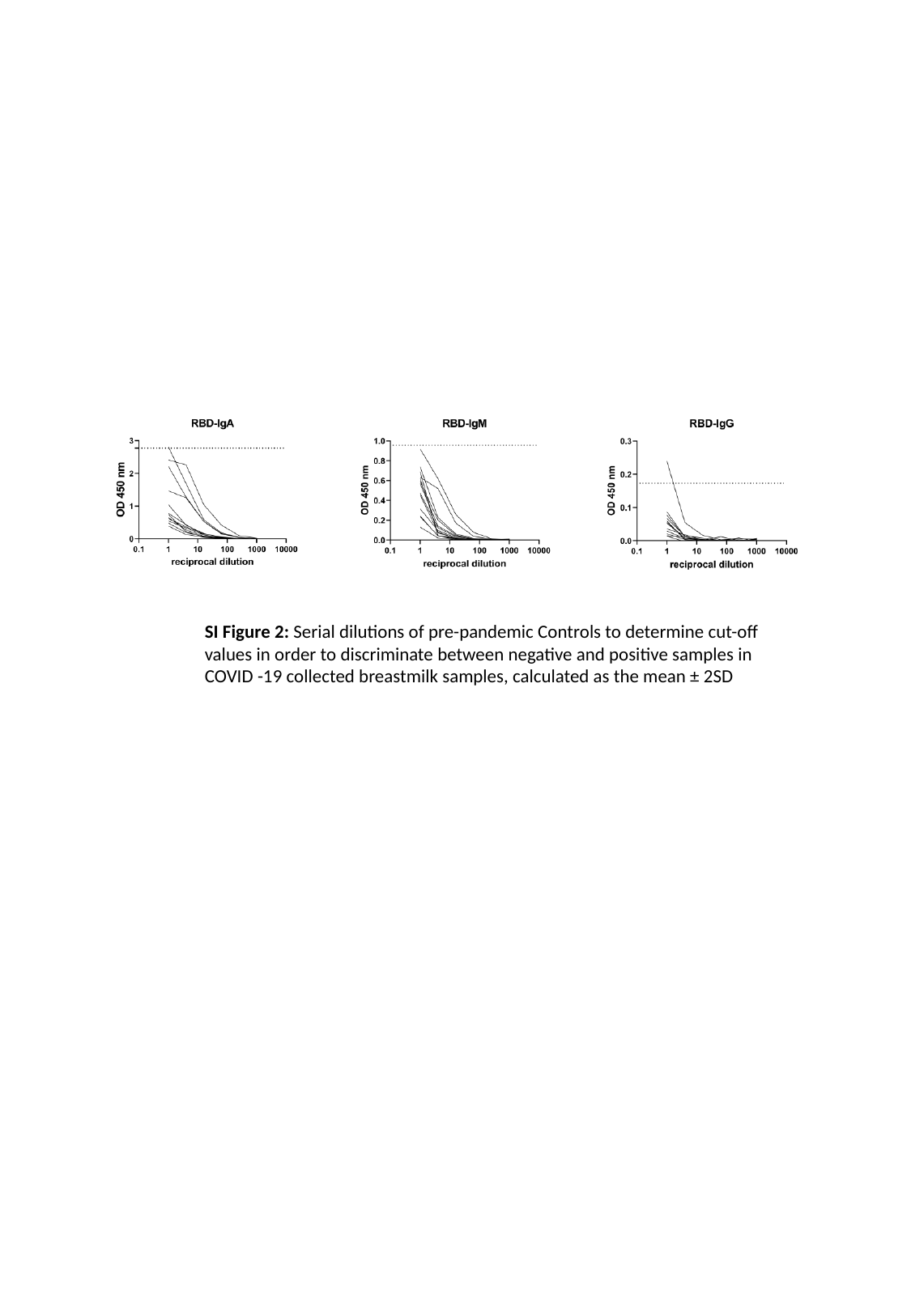

SI Figure 2: Serial dilutions of pre-pandemic Controls to determine cut-off values in order to discriminate between negative and positive samples in COVID -19 collected breastmilk samples, calculated as the mean ± 2SD

### Slide 3
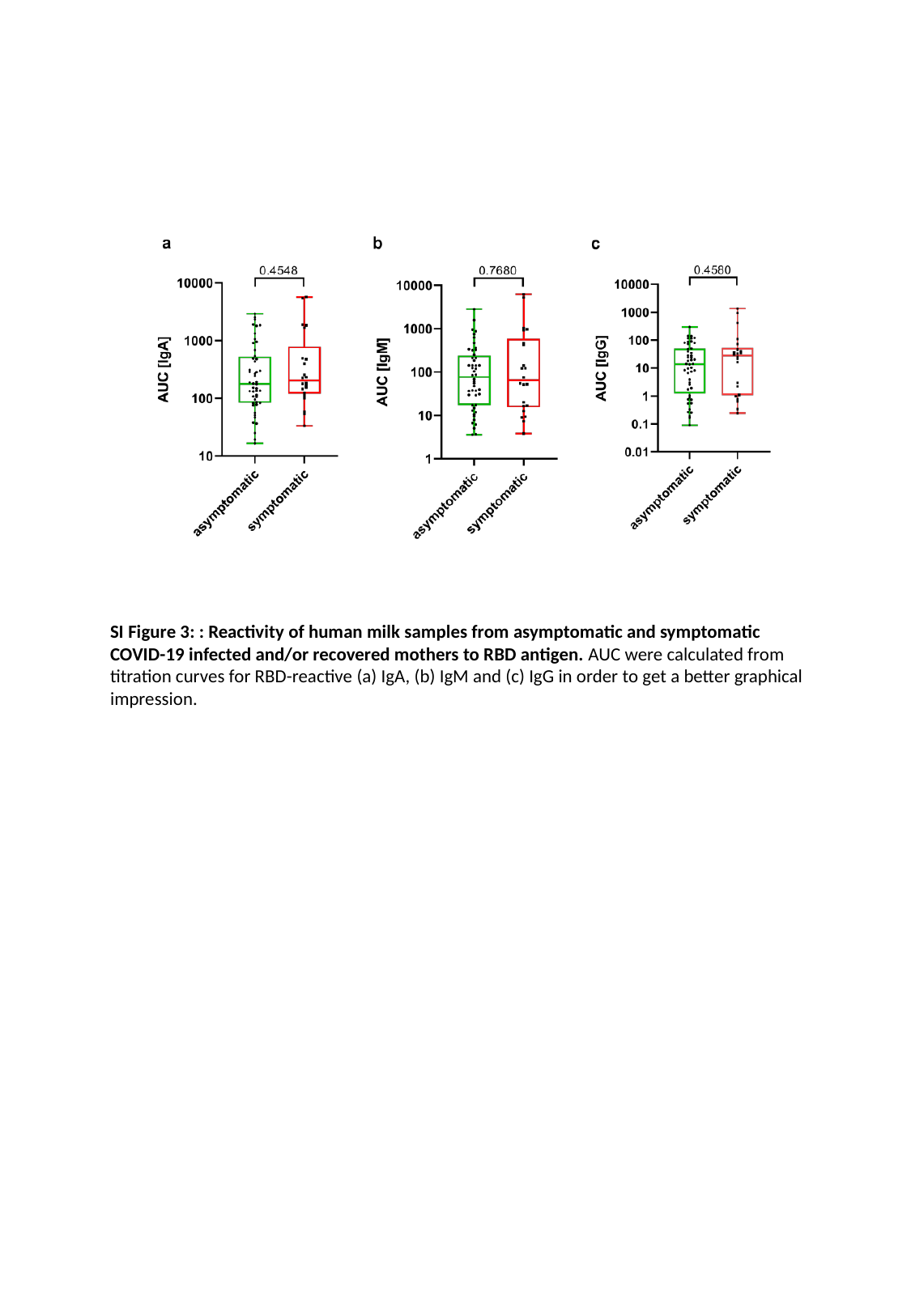

SI Figure 3: : Reactivity of human milk samples from asymptomatic and symptomatic COVID-19 infected and/or recovered mothers to RBD antigen. AUC were calculated from titration curves for RBD-reactive (a) IgA, (b) IgM and (c) IgG in order to get a better graphical impression.
